# WHO early warning indicators of HIV drug resistance in children and adolescents living with HIV in Cameroon: a pre- and post-COVID-19 analysis

**DOI:** 10.64898/2026.02.10.26346049

**Authors:** Armando B. D. Djiyou, Carole Else Eboumbou Moukoko, Palmer M. Netongo, Niels Kaze, Bernard Fortune Melingui, Ide Armelle Djuidje Chatue, Yoann Madec, Avelin F. Aghokeng, Calixte Ida Penda

## Abstract

COVID-19 disrupted global health service delivery, particularly among children and adolescents living with HIV (CALHIV), increasing the risk of poor treatment adherence. This study assessed the impact of the COVID-19 pandemic on WHO-recommended early warning indicators (EWIs) of HIV drug resistance (HIVDR) among CALHIV. We conducted a descriptive, longitudinal, retrospective study among children (0–9 years) and adolescents (10–19 years) receiving antiretroviral therapy (ART) in five health facilities in the Littoral region of Cameroon. Seven EWIs were monitored: ART attrition, viral load (VL) suppression, VL coverage, appropriate second VL, ARV medicine stock-outs, antiretroviral drug refills at the pharmacy, and appropriate switch to second-line ART. EWI were collected from January 2018 to December 2021 and classified as “poor,” “fair,” or “desirable” according to WHO criteria. Trend analyses were performed using Pearson’s Chi-squared test with Yates’ correction in R (version 4.1.1). In 2021, 817 participants were included, comprising 214 children and 603 adolescents. Overall performance was poor for most EWIs in both age groups, except for ART attrition and VL coverage, which showed desirable performance across years. A slight improvement in most indicators was observed between 2018 and 2019, followed by a significant decline in 2020 (p<0.001), coinciding with strict COVID-19 restrictions, and a subsequent improvement in 2021 (p<0.01) as mitigation measures were eased. Despite this recovery, children consistently experienced worse outcomes, including higher ART attrition (9.4% vs 4.4%, p<0.05), lower VL suppression (75.3% vs 82.1%, p<0.05), and poorer access to confirmatory VL testing (15.1% vs 69.5%, p<0.001). Overall, the COVID-19 pandemic negatively affected HIV service delivery during its early phase, although the health system demonstrated adaptive capacity one year later. Targeted public health actions are therefore needed to prevent their long-term effects and improve treatment outcomes in this vulnerable population, especially among children.

## Introduction

According to the United Nations Children’s Fund (UNICEF), around 2.38 million children and adolescents aged 0–19 years were living with HIV in 2023 [1]. Despite overall gains in the global HIV response, children and adolescents living with HIV (CALHIV) continue to fall behind adults in HIV testing, antiretroviral therapy (ART) initiation, retention, and viral suppression. The rapid scale-up of ART, coupled with the “test and treat” strategy, has significantly reduced HIV-related morbidity and mortality. However, the positive impact of this scale-up is increasingly threatened by the growing prevalence of both transmitted and acquired HIV drug resistance (HIVDR) in resource-limited settings (RLS) [2].

Given that HIVDR testing is not routinely available nor recommended in RLS such as Cameroon, the World Health Organization (WHO) has released a comprehensive HIVDR surveillance strategy [3,4], which was followed in Africa by a regional action plan to prevent and respond to HIVDR [5]. Aside from surveillance of pre-treatment and acquired drug resistance, this strategy strongly promotes the routine assessment of early warning indicators (EWI) as an alternative and robust method to evaluate the risk of HIVDR emergence [6]. EWIs are ART site factors that may be associated with preventable emergence of HIVDR, and can be acted on at the ART site or program level [5]. According to WHO guidelines, EWI assessments should be conducted annually using routinely collected data from patient medical and pharmacy records [3]. However, nationwide annual reporting of EWI in RLS is often hindered by financial and logistical constraints. As an alternative, many countries have adopted facility-based EWI monitoring to support more targeted surveillance efforts. Indeed, previous assessments of EWIs conducted both at the national [7–12] and facility [13–19] levels in several African countries have been instrumental in improving the delivery of HIV services and reducing the emergence and transmission of HIVDR in those countries.

In Cameroon, five rounds of EWI monitoring have been conducted since 2008, leading to the development and implementation of national- and clinic-level recommendations after each round based on the findings [5]. Nevertheless, while the WHO recommends that EWI should be monitored separately, even if both pediatric and adult patients are treated at the same clinic, most of those studies have been focused on adults or have not disaggregated data by age group. To our knowledge, only one study assessed EWI among adolescents receiving ART in Cameroon, but this study was conducted before 2020, a pivotal year marked by the onset of the COVID-19 pandemic. Yet, many reports highlighted that the COVID-19 pandemic significantly disrupted health service delivery, including HIV care, with widespread stay-at-home orders and ART stock-outs limiting patient access to care [20,21]. In 2020, Cameroon’s health ministry reported that of the 300,000 HIV-positive people in the country who need antiretrovirals, about 60% refused to visit hospitals because of COVID-19 [22]. Moreover, in a study conducted by our group between February and September 2021, we showed that up to 27.2% (76/280) of adolescents on ART in a clinic hosting the reference viral load (VL) laboratory in the Littoral region of Cameroon did not receive a VL test during the last 12 months [23]. This suboptimal delivery of HIV care and treatment is particularly concerning because it happened in an era of transition to dolutegravir (DTG)-based regimens [2], as it may accelerate the emergence of drug-resistant viruses, which could impair the long-term effectiveness of these new regimens at the population level.

The aim of our study was to evaluate EWIs of HIVDR in CALHIV before and during the COVID-19 pandemic in order to identify the determining factors of poor therapeutic compliance.

## Methods

### Study design, settings, and target population

A descriptive, longitudinal, and retrospective study was conducted between January and June 2022 in Cameroon. During the study period, EWI of the last four years (January 2018 to December 2021) were monitored among CALHIV aged 0 – 19 years, and receiving ART in five urban health facilities located in Douala, the Littoral Region of Cameroon: Laquintinie Hospital of Douala, Nylon District Hospital, *Cité des Palmiers* District Hospital, New-Bell District Hospital, and *Centre Médical d’Arrondissement* de Soboum (CMA Soboum). The aforementioned health facilities were selected based on their extensive experience in pediatric ART management and the availability of first- and second-line ART regimens at the hospital pharmacy, as well as ART registers and patient medical files. The source population consisted of children (0-9 years) and adolescents (10-19 years) enrolled and/or followed up at the selected study sites annually.

### Selection of early warning indicators

EWI was selected based on relevance to anticipated program interventions and the availability of data at the health facility. Using the most updated set of standard quality-of-care indicators of HIVDR included in the 2020 WHO consolidated HIV strategic information guidelines [24], seven EWI were monitored: EWI-1 ‘*total attrition from ART*’ (a proxy measure of retention), EWI-2 ‘*antiretroviral drug refills at the pharmacy*’ (a proxy measure of appropriate adherence to ART), EWI-3 ‘*viral load testing coverage*’ (assesses the extent to which VL testing is available in the facility and enables appropriate interpretation of VL testing data), EWI-4 ‘*people living with HIV who have suppressed viral load*’ (measures clinical outcomes of patients on ART regardless of initiation date), EWI-5 ‘*appropriate second viral load test*’ (measures the extent to which people living with HIV with non-suppressed VL receive appropriate follow-up VL testing to check virologic suppression), EWI-6 ‘*appropriate switch to second-line ART*’ (proxy measure on how well a facility uses viral load testing results to identify failure and promptly switch people to the next line of treatment), and EWI-7 ‘*antiretroviral medicine stock-out*’. Table 1 provides definitions of the numerator and denominator for each indicator and their respective performance targets.

**Table 1:** Definition of WHO-recommended EWIs of HIV drug resistance and their respective performance targets.

| EWI and title | Description | Numerator | Denominator | Performance strata |
| --- | --- | --- | --- | --- |
| EWI-1: Total attrition from antiretroviral treatment (ART) | Number and percentage of people living with HIV reported on ART at the end of the last reporting period and/or newly initiating ART during the current reporting period who were not on ART at the end of the reporting period | The number of people living with HIV on ART at the end of the previous reporting period, plus the number of people living with HIV started on ART during the current reporting period, who are classified as having died, stopped treatment and/or been lost to follow-up by the end of the current period | The number of people living with HIV who are on ART at the end of the previous reporting period, plus the number of people living with HIV newly initiated on ART during the reporting period | Desirable performance (green): <15%<br>Fair performance (amber): 15–25%<br>Poor performance (red): >25% |
| EWI-2: ART adherence proxy (ARV refills) | Percentage of people receiving ART who pick up all prescribed ARV drugs on time (no more than two days late at the first drug pick-up after a defined baseline pick-up) | Number of patients who pick up all prescribed ARV drugs no more than two days late at the first pick-up after a defined baseline pick-up | Number of patients who picked up ARV drugs on or after the designated ART start date | Desirable performance (green): >90%<br>Fair performance (amber): 80–90%<br>Poor performance (red): <80% |
| EWI-3: VL testing coverage | Percentage of people on ART (for at least 6 months) with VL test results | Number of ART patients with at least one routine VL test result during the reporting period | Number of ART patients on ART at least 6 months | Desirable performance (green): >95%<br>Fair performance (amber): 85–95%<br>Poor performance (red): <85% |
| EWI-4: People living with HIV who have suppressed viral load (VL) | Percentage of people living with HIV receiving ART (for at least six months) who have suppressed VL (defined as VL <1000 copies/mL) during the reporting period | Number of people living with HIV on ART for at least 6 months and with at least one routine VL test result who have virological suppression (<1000 copies/mL) during the reporting period | Number of people living with HIV on ART at least 6 months, with at least one routine VL result in a medical or lab record during the reporting period | Desirable performance (green): >90%<br>Fair performance (amber): 80–90%<br>Poor performance (red): <80% |
| EWI-5: Appropriate second VL test | Percentage of people receiving ART with VL ≥1000 copies/mL who received a follow-up viral load test within six months after enhanced adherence counselling | Number of people living with HIV on ART who received a follow-up VL test within 6 months after a VL test result of ≥1000 copies/ml during the reporting period | Number of people living with HIV on ART with VL ≥1000 copies/ml during the reporting period. | Desirable performance (green): ≥90%<br>Poor performance (red): <90% |
| EWI-6: Appropriate switch to second-line ART | Percentage of patients with confirmed failure to suppress VL who switch to second-line ART within 90 days | Number of people on ART with confirmatory VL ≥1000 copies/mL who are switched to second-line ART within 90 days of the confirmatory VL test result of ≥1000 copies/mL | Total number of people on ART with confirmatory VL test result >1000 copies/mL | Desirable performance (green): 100%<br>Poor performance (red): <100% |
| EWI-7: Antiretroviral (ARV) medicine stock-out | Percentage of months with any day(s) of stock-out of any routinely dispensed ARV drug during the reporting period (12 months) | Number of months in the designated year in which there were stock-out days of any ARV drug routinely used at the site | 12 months of the reporting period | Desirable performance (green): 0%<br>Poor performance (red): >0% |

### Data abstraction and quality control

During 6 months, data from January 2018 to December 2021 were collected from ART registers, patient medical files, and pharmacy records, and then retrieved using a modified version of the standardized Excel-based WHO pre-established data collection tools [25]. This modified version of the data abstraction tool was tailored to capture the updated list of EWIs and some socio-demographic (sex, age) and clinical (current and previous ART regimens) characteristics. For nationwide assessment of EWI, WHO recommends data abstraction on a minimum number of consecutive patients at each facility to estimate EWI and achieve a 95% confidence interval of ±7% [25]. However, in this study, medical files of CALHIV receiving ART at each facility were systematically sampled until full sampling during the reporting period.

To ensure reliability, data was collected by two independent investigators (PhD students) trained on HIV data management using the modified EXCEL tools and then cross-checked for consistency. Incoherent data was resolved by retrieving additional source documents available at the health facility. Two senior researchers with experience in assessing EWI supervised the data collection process to ensure that the process is in line with the approved protocol and up to the ethical standard.

### Data analysis

Frequencies and proportions using all available data were calculated for each EWI overall, and separately for each facility, and were then stratified for children and adolescent populations. EWIs were classified as poor, fair, and desirable performance following the WHO-recommended thresholds. For trend analysis, statistical significance over time was analyzed for the performance of each EWI using the Pearson’s Chi-squared test with Yates’ continuity correction. Statistical analysis was done using R (version-4.1.1), and p-values < 0.05 were considered statistically significant.

### Ethical consideration

Ethical approval for this study was granted by the Institutional Ethics Committee for Research on Human Health of the University of Douala (N°2933/IEC-UD/10/2021/M). Administrative authorizations were obtained from the Littoral Regional Delegation of the Ministry of Public Health (N°1325/AAR/MINSANTE/DRSPL/BCASS) and each of the health facilities involved in the study. Confidentiality was respected during data abstraction by the use of a specific identification code for each enrolled patient number, and no personal identifiable information of the patients was collected. Since the study was done retrospectively, no contact with patients occurred during this study. Data were collected from medical/ART and pharmacy registers available at the selected study sites. Thus, informed consent from participants was not required.

## Results

### Baseline characteristics of the participants

Overall, 739 medical files from CALHIV receiving ART at the selected study sites were included in 2018, 813 in 2019, 832 in 2020, and 817 in 2021, with more than half of them followed up at the Laquintinie Hospital of Douala. Disaggregated by age group, the number of adolescents progressively increased from 425/739 (57.5%) in 2018 to 603/817 (73.8%) in 2021, while the number of children progressively decreased from 314 (42.5%) in 2018 to 214 (26.2%) in 2021. Across the years, the sex ratio was almost 1:1, as female participants accounted for ∼51% of the study participants each year. In this study, 8 out of 10 participants received Nevirapine- or Efavirenz (EFV)-based treatment as initial treatment. Regarding the current ART, we observed a decreasing trend in the number of CALHIV receiving EFV-based treatment, from 501 (70.0%) in 2018 to 174 (22.8%) in 2021; meanwhile, the number of CALHIV receiving DTG-based treatment increased from 1 (0.1%) to 324 (42.5%) during the same period (Table 2).

**Table 2.** Baseline characteristics of the study population.

|  | Year 2018<br>(N = 739) | Year 2019<br>(N = 813) | Year 2020<br>(N = 832) | Year 2021<br>(N = 817) |
| --- | --- | --- | --- | --- |
| Study sites |  |  |  |  |
| Laquintinie Hospital Douala | 425 (57.5%) | 447 (55.0%) | 418 (50.2%) | 380 (46.5%) |
| Nylon District Hospital | 108 (14.6%) | 128 (15.7%) | 148 (17.8%) | 156 (19.1%) |
| *Cité des Palmiers* District Hospital | 83 (11.2%) | 101 (12.4%) | 115 (13.8%) | 114 (14.0%) |
| New-Bell District Hospital | 49 (6.6%) | 54 (6.6%) | 66 (7.9%) | 80 (9.8%) |
| CMA Soboum | 74 (10.0%) | 83 (10.2%) | 85 (10.2%) | 87 (10.6%) |
| Age category |  |  |  |  |
| Children (0-9 years) | 314 (42.5%) | 302 (37.1%) | 262 (31.5%) | 214 (26.2%) |
| Adolescents (10-19 years) | 425 (57.5%) | 511 (62.9%) | 570 (68.5%) | 603 (73.8%) |
| Gender |  |  |  |  |
| Female | 377 (51.0%) | 413 (50.8%) | 426 (51.2%) | 417 (51.0%) |
| Male | 362 (49.0%) | 400 (49.2%) | 406 (48.8%) | 400 (49.0%) |
| ART history |  |  |  |  |
| ART at initiation |  |  |  |  |
| EFV-based | 294 (39.8%) | 344 (42.3%) | 363 (43.6%) | 357 (43.7%) |
| NVP-based | 330 (44.7%) | 335 (41.2%) | 325 (39.1%) | 310 (37.9%) |
| LPV/r-based | 115 (15.6%) | 134 (16.5%) | 144 (17.3%) | 144 (17.6%) |
| DTG-based | 0 | 0 | 0 | 6 (0.7%) |
| Currently on ART |  |  |  |  |
| EFV-based | 501 (70.0%) | 512 (68.5%) | 499 (65.1%) | 174 (22.8%) |
| NVP-based | 19 (2.7%) | 7 (0.9%) | 0 | 0 |
| DTG-based | 1 (0.1%) | 1 (0.1%) | 24 (3.1%) | 324 (42.5%) |
| LPV/r-based | 184 (25.7%) | 208 (27.8%) | 206 (26.9%) | 159 (20.9%) |
| ATV/r-based | 11 (1.5%) | 19 (2.5%) | 38 (5.0%) | 105 (13.8%) |

ART, antiretroviral treatment; ATV/r, Atazanavir boosted with ritonavir; CMA Soboum, *Centre Médical* *d'Arrondissement de Soboum*; DTG, Dolutegravir; EFV, Efavirenz; LPV/r, Lopinavir boosted with ritonavir; NVP, Nevirapine;

### Performance of each EWI

Table 3 presents the overall and age-specific performance for each EWI.

**Table 3.** Summary of EWI performance stratified by age category and year.

|  |  | EWI-1: Total attrition from ART | EWI-2: ARV refills at the pharmacy | EWI-3: VL testing coverage | EWI-4: PLHIV who have a suppressed VL | EWI-5: Appropriate second VL test | EWI-6: Appropriate switch 2<sup>nd</sup> line | EWI-7: ARV medicine stock-out |
| --- | --- | --- | --- | --- | --- | --- | --- | --- |
| Children<br>(0-9 years) | Year 2018 | 5.7%<br>(18/314) | 77.3%<br>(222/287) | 89.1%<br>(253/284) | 66.7%<br>(166/249) | 48.2%<br>(40/83) | 26.9%<br>(7/26) | 100%<br>(12/12) |
|  | Year 2019 | 11.7%<br>(35/299) | 72.1%<br>(181/251) | 92.6%<br>(224/242) | 81.7%<br>(183/224) | 43.9%<br>(18/41) | 22.2%<br>(2/9) | 100%<br>(12/12) |
|  | Year 2020 | 8.1%<br>(21/259) | 56.6%<br>(129/228) | 78.2%<br>(176/225) | 77.7%<br>(136/175) | 53.8%<br>(21/39) | 41.2%<br>(7/17) | 100%<br>(12/12) |
|  | Year 2021 | 9.4%<br>(20/212) | 78.4%<br>(145/185) | 88.1%<br>(163/185) | 75.3%<br>(122/162) | 15.1%<br>(5/33) | 100%<br>(2/2) | 100%<br>(12/12) |
| Adolescents<br>(10-19 years) | Year 2018 | 1.2%<br>(5/425) | 81.5%<br>(340/417) | 92.7%<br>(383/413) | 71.5%<br>(274/383) | 54.1%<br>(59/109) | 12.8%<br>(5/39) | 100%<br>(12/12) |
|  | Year 2019 | 5.5%<br>(28/509) | 81.9%<br>(388/474) | 91.5%<br>(430/470) | 77.0%<br>(331/430) | 47.8%<br>(47/99) | 57.1%<br>(16/28) | 100%<br>(12/12) |
|  | Year 2020 | 6.6%<br>(37/563) | 63.8%<br>(335/525) | 82.1%<br>(431/525) | 74.7%<br>(322/431) | 61.5%<br>(67/109) | 45.5%<br>(10/22) | 100%<br>(12/12) |
|  | Year 2021 | 4.4%<br>(26/594) | 79.6%<br>(442/555) | 92.6%<br>(514/555) | 82.1%<br>(422/514) | 69.5%<br>(57/82) | 100%<br>(6/6) | 100%<br>(12/12) |
| Overall | Year 2018 | 3.1%<br>(23/739) | 79.8%<br>(562/704) | 91.2%<br>(636/697) | 69.6%<br>(440/632) | 51.6%<br>(99/192) | 18.5%<br>(12/65) | 100%<br>(12/12) |
|  | Year 2019 | 7.8%<br>(63/808) | 78.5%<br>(569/725) | 91.8%<br>(654/712) | 78.6%<br>(514/654) | 46.4%<br>(65/140) | 48.6%<br>(18/37) | 100%<br>(12/12) |
|  | Year 2020 | 7.1%<br>(58/822) | 61.6%<br>(464/753) | 81.0%<br>(607/750) | 75.6%<br>(458/606) | 59.5%<br>(88/148) | 43.6%<br>(17/39) | 100%<br>(12/12) |
|  | Year 2021 | 5.7%<br>(46/806) | 79.3%<br>(587/740) | 91.5%<br>(677/740) | 80.5%<br>(544/676) | 53.9%<br>(62/115) | 100.0%<br>(8/8) | 100%<br>(12/12) |

ART, antiretroviral treatment; ARV, antiretroviral; VL, viral load.

For EWI-1 (*total attrition from ART*), the overall performance was consistently excellent across all the rounds of EWI surveys from 2018 to 2021, as less than 15% of loss from ART was reported each year. Disaggregated data still showed good performances in children and adolescents, although children tended to have a higher rate of loss from ART compared to adolescents in 2018 (5.7% vs. 1.2%, p<0.001), 2019 (11.7% vs. 5.5%, p=0.001), and 2021 (9.4% vs. 4.4%, p=0.006).

For EWI-2 (*antiretroviral drug refills at the pharmacy*), the overall performance was consistently poor from 2018 to 2021, as less than 80% of the CALHIV picked up their prescribed ARV drugs on time each year. Disaggregated data showed that while children maintained poor performance across the year, adolescents reached a fair level of performance (between 80-90%) in 2018 (77.3% vs. 81.5%, p=0.2), and 2019 (72.1% vs. 81.9%, p=0.002).

For EWI-3 (*viral load testing coverage*), the overall performance was fair (between 85%-95%) in 2018, 2019, and 2021, but poor (<85%) in 2020. The same trend was observed when stratified for children and adolescents, but children had a lower rate of VL coverage in 2018 (89.1% vs. 92.7%, p=0.06), and 2021 (88.1% vs. 92.6%, p=0.04).

For EWI-4 (*people living with HIV who have suppressed viral load*), the target of ≥90% of VL suppression was missed across the year. The overall performance was poor (<80%) from 2018 to 2020 and fair (between 80%-90%) in 2021. When stratified according to age groups, a fair performance was reached in 2019 for children and in 2021 for adolescents. However, a statistically significant difference between those two age groups was only found in 2021 (75.3% vs. 82.1%, p=0.04).

For EWI-5 (*appropriate second viral load test*), the overall performance was consistently poor from 2018 to 2021, as less than 90% of the CALHIV receiving ART with a VL ≥1000 copies/mL had a follow-up VL test within six months after enhanced adherence counselling each year. Disaggregated data still showed a poor performance in both children and adolescents, but children had a lower percentage of follow-up VL compared to adolescents in 2021 (15.1% vs. 69.5%, p<0.001).

For EWI-6 (*appropriate switch to second-line ART*), the target of switching to second-line all CALHIV with confirmed failure to suppress viral loads was achieved only in 2021. The overall performance was poor from 2018 to 2020. A similar trend was observed in disaggregated data, and no statistically significant difference was found between children and adolescents in any of the reporting years.

For EWI-7 (*antiretroviral medicine stock-out*), the overall and disaggregated performances were poor across the years, as we found disruption of at least one of the dispensed ARVs per month during the reporting period.

### Performance of each EWI according to the study sites

Reported to the study sites (S1 Table), all the health facilities met the WHO target of total attrition from ART <15% each year. However, none of them met the target of achieving follow-up VL testing ≥90% within six months of enhanced adherence counselling, nor the target of no month with any day(s) of stock out of any routinely dispensed ARV drug during the 12-month reporting period. Regarding the other EWI, although the performances were globally poor, we found variation according to study sites. Notably, 2/5 (40.0%) achieved VL testing coverage ≥95% in 2018 (Laquintinie Hospital of Douala and Cité des Palmiers District Hospital) and 2021 (Laquintinie Hospital of Douala and Nylon District Hospital). The target of ≥90% of individuals who picked up their prescribed ARV drugs was achieved by the Cité des Palmiers District Hospital each of the reported years and by the Nylon District Hospital in 2018, 2019, and almost in 2021. All the health facilities performed well on switching eligible CALHIV to second-line regimens in 2021. A summary of health facility performance is available in Table 4.

**Table 4.** Overall EWI facility performance according to WHO targets (n =5).

|  | Desirable |  |  |  | Fair |  |  |  | Poor |  |  |  |
| --- | --- | --- | --- | --- | --- | --- | --- | --- | --- | --- | --- | --- |
|  | 2018 | 2019 | 2020 | 2021 | 2018 | 2019 | 2020 | 2021 | 2018 | 2019 | 2020 | 2021 |
| EWI-1:<br>Total attrition<br>from ART | 5/5<br>(100.0%) | 5/5<br>(100.0%) | 5/5<br>(100.0%) | 5/5<br>(100.0%) | 0/5<br>(0.0%) | 0/5<br>(0.0%) | 0/5<br>(0.0%) | 0/5<br>(0.0%) | 0/5<br>(0.0%) | 0/5<br>(0.0%) | 0/5<br>(0.0%) | 0/5<br>(0.0%) |
| EWI-2:<br>ARV refills at the<br>pharmacy | 2/5<br>(40.0%) | 2/5<br>(40.0%) | 1/5<br>(20.0%) | 1/5<br>(20.0%) | 0/5<br>(0.0%) | 0/5<br>(0.0%) | 0/5<br>(0.0%) | 2/5<br>(40.0%) | 3/5<br>(60.0%) | 3/5<br>(60.0%) | 4/5<br>(80.0%) | 2/5<br>(40.0%) |
| EWI-3:<br>VL testing<br>coverage | 2/5<br>(40.0%) | 0/5<br>(0.0%) | 0/5<br>(0.0%) | 2/5<br>(40.0%) | 1/5<br>(20.0%) | 5/5<br>(100.0%) | 2/5<br>(40.0%) | 2/5<br>(40.0%) | 2/5<br>(40.0%) | 0/5<br>(0.0%) | 3/5<br>(60.0%) | 1/5<br>(20.0%) |
| EWI-4:<br>PLHIV who have a<br>suppressed VL | 0/5<br>(0.0%) | 0/5<br>(0.0%) | 0/5<br>(0.0%) | 0/5<br>(0.0%) | 1/5<br>(20.0%) | 2/5<br>(40.0%) | 0/5<br>(0.0%) | 3/5<br>(60.0%) | 4/5<br>(80.0%) | 3/5<br>(60.0%) | 5/5<br>(100.0%) | 2/5<br>(40.0%) |
| EWI-5:<br>Appropriate<br>second VL test | 0/5<br>(0.0%) | 0/5<br>(0.0%) | 0/5<br>(0.0%) | 0/5<br>(0.0%) | N/A | N/A | N/A | N/A | 5/5<br>(100.0%) | 5/5<br>(100.0%) | 5/5<br>(100.0%) | 5/5<br>(100.0%) |
| EWI-6:<br>Appropriate switch<br>to second-line ART | 0/5<br>(0.0%) | 1/5<br>(20.0%) | 0/5<br>(0.0%) | 5/5<br>(100.0%) | N/A | N/A | N/A | N/A | 5/5<br>(100.0%) | 4/5<br>(80.0%) | 5/5<br>(100.0%) | 0/5<br>(0.0%) |
| EWI-7:<br>ARV medicine<br>stock-out | 0/5<br>(0.0%) | 0/5<br>(0.0%) | 0/5<br>(0.0%) | 0/5<br>(0.0%) | N/A | N/A | N/A | N/A | 5/5<br>(100.0%) | 5/5<br>(100.0%) | 5/5<br>(100.0%) | 5/5<br>(100.0%) |

### EWI trend analysis

Fig 1 presents the trend of each EWI across the four reporting years.

**Fig 1.**
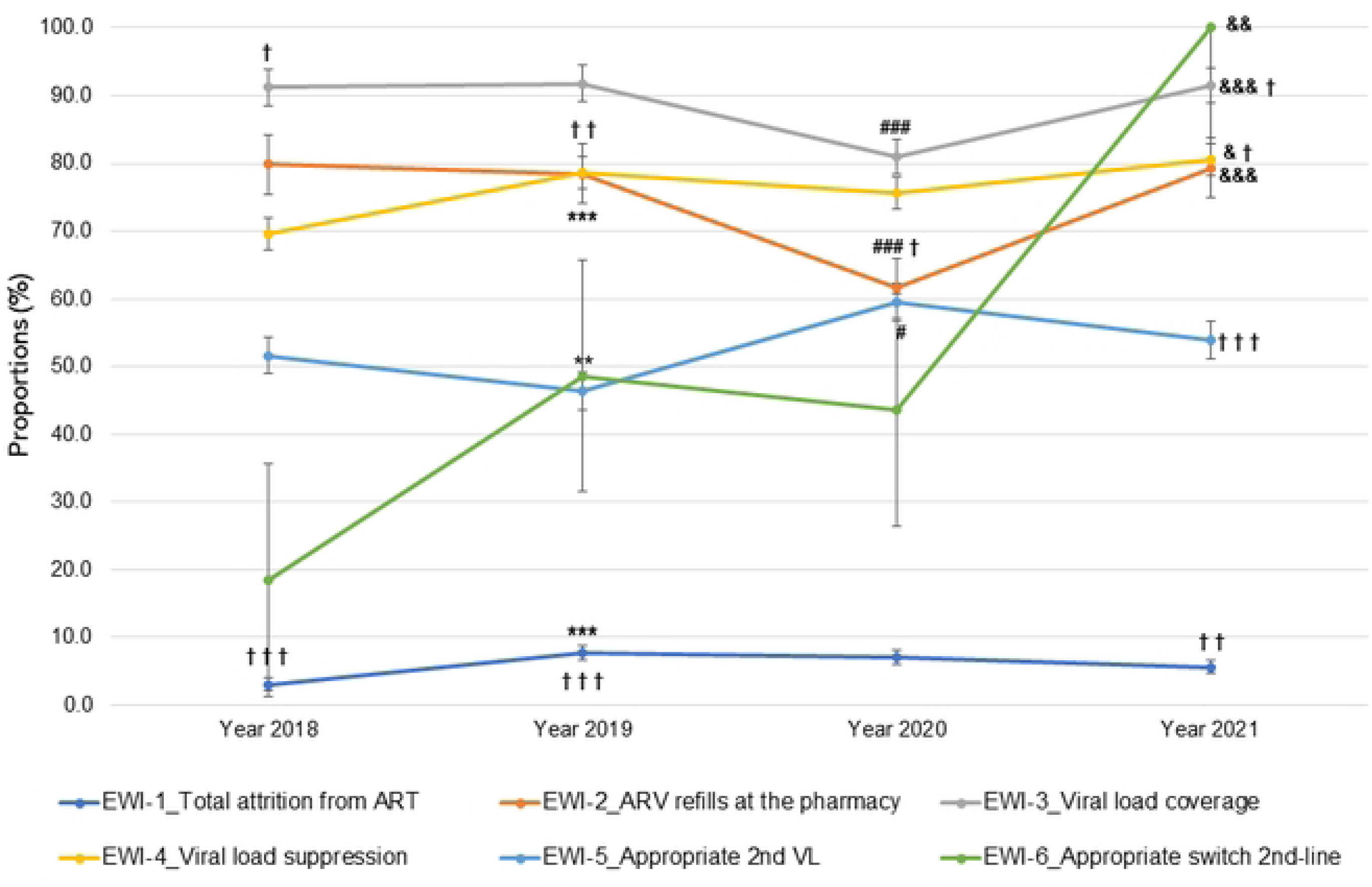
Trend of early warning indicators at the selected study sites between 2018 and 2021. The figure illustrates the overall trends of 6 out of the 7 WHO-recommended early warning indicators of HIV drug resistance during the reporting period (January 2018 to December 2021). The chi-square test, along with Yates’ continuity correction, was used to compare performance between years. The symbol ’*’ indicates a significant difference in performance between 2018 and 2019, the symbol ‘#’ indicates a significant difference between 2019 and 2020, the symbol ‘&’ indicates a significant difference between 2020 and 2021, and the symbol ‘†’ indicates a significant difference between children (0-9 years) and adolescents (10-19 years). One symbol denotes a p-value <0.05, two symbols indicate p <0.01, and three symbols indicate p <0.001.

Overall, between 2018 and 2019, we found an increasing trend for 4 out of 6 EWI, with three of them found statistically significant, namely EWI-1 ‘*total attrition from ART*’ (3.1% vs. 7.8%, P<0.001), EWI-4 ‘*people living with HIV who have suppressed viral load*’ (69.6% vs. 78.6%, p<0.001), and EWI-6 ‘*appropriate switch to second-line ART*’ (18.5% vs. 48.6%, p=0.003). The two others, EWI-2 ‘*antiretroviral drug refills at the pharmacy*’ and EWI-5 ‘*appropriate second viral load test*’ had a decreasing trend, even though not statistically significant.

Between 2019 and 2020, apart from EWI-5 ‘*appropriate second viral load test*’ that significantly increased (46.4% vs. 59.5%, p=0.03), a decreasing trend was observed for the 5 other EWI, with statistically significant differences found for EWI-2 ‘*antiretroviral drug refills at the pharmacy*’ (78.5% vs. 61.6%, p<0.001) and EWI-3 ‘*viral load testing coverage*’ (91.8% vs. 81.0%, p<0.001).

Between 2020 and 2021, the overall performance of 4 out of 6 EWI significantly increased, namely EWI-2 ‘*antiretroviral drug refills at the pharmacy*’ (61.6% vs. 79.3%, p<0.001), EWI-3 ‘*viral load testing coverage*’ (81.0% vs. 91.5%, p<0.001), EWI-4 ‘*people living with HIV who have suppressed viral load*’ (75.6% vs. 80.5%, p<0.04) and EWI-6 ‘*appropriate switch to second-line ART*’ (43.6% vs. 100.0%, p=0.01). There was a drop in EWI-1 ‘*total attrition from ART*’ and EWI-5 ‘*appropriate second viral load test*’ that was not shown to be significant.

## Discussion

Annual monitoring of EWIs allows national HIV/AIDS control programs in RLS to assess the quality of HIV service delivery offered to HIV-infected people on ART and propose corrective measures to improve ART response [4]. Our results show that EWI performance has been globally poor in recent years in the Littoral Region of Cameroon. Access to routine health services was disrupted for CALHIV, particularly in 2020 due to the strict restrictions implemented during the COVID-19 pandemic, followed by an adaptation of the response in 2021 when these restrictions were gradually lifted.

In our study population, we observed a declining trend in the number of children followed up at the selected health facilities, while the number of adolescents progressively increased. This reflects the success of prevention of mother-to-child transmission (PMTCT) programs and improved access to treatment and care, which have contributed to a greater number of children living with HIV surviving into adolescence [1]. Additionally, we observed a shift in the proportion of CALHIV receiving EFV-based regimens toward DTG-based regimens, a trend that was anticipated considering the latest WHO guidelines recommending the transition to DTG as the preferred option for both first- and second-line ART in RLS [26].

While retention has long been a major concern in many RLS including Cameroon [7–10,12,13], we found a satisfactory ART retention rate in our study, with performance consistently exceeding the WHO threshold of 85% throughout the reporting period. These results are encouraging as they reflect the successful implementation of several programmatic interventions in recent years to ensure continuous access to ART and close follow-up of CALHIV [13]. Key interventions included the recruitment and training of social workers who provide adherence counseling and facilitate follow-up for patients who miss appointments [9], implementation of differentiated service delivery models tailored to the needs of CALHIV such as adolescent-friendly clinics and school-based care, and peer-led initiatives including “adolescent champions” and support groups [27,28]. Collectively, these strategies likely contributed to the improved ART retention among CALHIV observed in our study.

In contrast, the performance of the other early warning indicators remained suboptimal throughout the study period and varied across health facilities. Interestingly, peripheral health facilities outperformed the central-level health facility, Laquintinie Hospital of Douala, in some of those key indicators. For example, both *Cité des Palmiers* and Nylon District Hospitals met the 90% target for timely ART pickup among CALHIV, while CMA Soboum was the only facility that almost achieved the 90% VL suppression target. These findings align with previous observations by Penda et al. [29], who reported significantly lower viral suppression rates among adolescents receiving care in higher-tier health facilities in Cameroon. A poor performance of EWIs has been previously reported in Cameroon [8–10,12] and other RLS [6,13–15,18,19]. In most of these studies, poor performance was attributed to programmatic challenges, suboptimal implementation of guidelines by healthcare providers, and patient-related factors. Frequent ART stock-outs and low VL testing coverage were likely due to logistical challenges and dysfunctions in the ARV and laboratory supply chains. Delays in switching to second-line ART were often related to the prolonged turnaround time for VL results (several months instead of the recommended 4 weeks [26]), as well as delayed prescription of VL by clinicians according to the national treatment monitoring algorithm [7].

Despite the already suboptimal performance observed in prior years, a comparative analysis revealed a declining trend of nearly all EWIs in 2020, a year marked by the onset of the COVID-19 pandemic. This might be explained by the widespread disruption of HIV service delivery due to tight COVID-19-related restriction measures, such as strict national lockdowns, fear of visiting hospitals, shutdowns of certain health facilities and pharmaceutical companies, redeployment of healthcare workers to COVID-19 response efforts, difficulties with international shipping due to border restrictions, and patients avoiding health facilities due to fear of contracting SARS-CoV-2 [30,31]. Due to these challenges, numerous CALHIV were unable to attend their routine clinical visits, access adherence counseling or therapeutic education sessions, or even receive ART and necessary VL monitoring. Early in the COVID-19 pandemic, the WHO sounded the alarm on this matter in a brief report, showing that COVID-19 led to reduced access to ART in 36 countries, which account for 45% of people on ART globally (approximately 11.5 million individuals), with 24 of them reporting inadequate ARV supply chains [32]. Additionally, in a systematic review and meta-analysis, nearly all included studies reported a significant decrease in ART availability and uptake, as well as in the number of scheduled visits during the pandemic [33]. This trend was also confirmed in Central Africa by a study involving 21 clinics across five countries in the region, which showed a significant reduction in access to ART, HIV testing and other laboratory services, especially VL testing [34].

Surprisingly, VL suppression and retention on ART were not significantly affected by the COVID-19 pandemic in our study. This might seem paradoxical, considering how essential uninterrupted access (and adherence) to ART, routine VL monitoring, and support programs are to achieving long-term viral suppression in CALHIV [35]. However, similar findings have been reported in both high-income countries [36] and resource-limited settings [37,38]. In Uganda, for example, while 41.8% of patients reported that the pandemic had affected their ability to attend clinic appointments, only 9.3% said they had experienced ARV shortages during lockdown, and just 8.3% reported poor adherence during that period [38]. This suggests that mitigation measures implemented by the country were effective, although long-term evaluations are still needed.

VL confirmation was the only EWI with an increasing trend in 2020, suggesting at first glance that it may not have been affected by the negative impacts of the COVID-19 pandemic. However, these results should be interpreted with caution. According to the national treatment algorithm, VL confirmation testing is recommended within 3 to 6 months following an initial unsuppressed result. Consequently, the significant decline observed in 2019 may, in fact, represent a delayed consequence of the VL testing disruptions encountered in 2020. Therefore, despite the increasing trend in 2020, it is likely that the pandemic impacted VL confirmation, though its effect was reflected during the previous reporting year.

A common feature across all these EWIs was a return by 2021, one year after the pandemic began, to performance levels comparable to or even better than those observed before the pandemic. In response to the COVID-19 pandemic, WHO and global partners such as UNAIDS, CDC/PEPFAR, and the Global Fund to Fight AIDS, Tuberculosis, and Malaria, accelerated the uptake of long-standing innovative interventions and strategies that had previously struggled to be scaled up [39,40]. The most reported interventions included multi-month dispensing (MMD) of ART, expansion of community service delivery, and the use of technology and virtual platforms for client engagement and site-level monitoring [37]. The implementation of these interventions helped national programs to mitigate the negative impacts on healthcare delivery, reversing the declining trend observed in 2020 [33]. Despite improvements, performance was worse for children compared to adolescents, as they had poorer attrition to ART (9.4% vs. 4.4%, p<0.05), lower VL suppression (75.3% vs. 82.1%, p<0.05), and inadequate access to VL testing with lower VL coverage (88.1% vs. 96.6%, p<0.05) and confirmatory VL performed (15.1% vs. 69.5%, p<0.001). These disparities may reflect challenges unique to younger children, such as greater dependence on caregivers, limited access to child-friendly formulations, and reduced healthcare engagement during the COVID-19 pandemic [21,37], highlighting the need for targeted, subpopulation-specific interventions to improve pediatric HIV outcomes in RLS.

This study provides a unique contribution by comprehensively assessing up-to-date quality-of-care indicators over a four-year period, including both pre- and post-pandemic periods. Nevertheless, it is important to acknowledge some limitations in our study. First, its retrospective design, which relies on routine programmatic data, may be subject to reporting biases, data incompleteness, and variability in data quality across facilities. Second, the study was conducted in a limited number of urban sites, which may not fully capture the heterogeneity of HIV care performance in semi-urban and rural settings. Despite these limitations, the findings provide valuable insights into programmatic gaps and how national programs have responded to COVID-19 disruptions. This is particularly relevant in the current context of reduced U.S. global health funding and emerging HIVDR to dolutegravir-based regimens across sub-Saharan Africa, as recently emphasized by the WHO [41]. Our study underscores the importance of strengthening health systems in RLS, optimizing VL monitoring, enhancing HIVDR surveillance to better inform ART strategies, and implementing targeted interventions to mitigate the risk of HIV drug resistance, especially among vulnerable subpopulations such as children and adolescents.

## Data Availability

All relevant data are within the paper and its supporting information files.

## Acknowledgements

We thank the Cameroon national health authorities and the participating study sites for granting the necessary research authorizations. We also express our sincere appreciation to the staff of these sites for their valuable support in providing the necessary records, information, and assistance throughout the data collection process.

## Supporting information

**S1 Table. Summary of site performance for each early warning indicator (EWI).** The table presents EWI performance, color-coded according to the WHO thresholds outlined in Table 1: green indicates desirable performance, amber indicates fair performance, and red indicates poor performance.

